# Machine learning to detect intraoperative ischemia from electroencephalography in carotid endarterectomy surgery

**DOI:** 10.64898/2026.08.01.26359458

**Authors:** Shyam Visweswaran, Mehdi Nourelahi, Amir I. Mina, Jessi U. Espino, Nihal Murali, Kayhan Batmanghelich, Parthasarathy D. Thirumala

## Abstract

Cerebral ischemia is a significant concern during high-risk surgeries, such as carotid endarterectomy (CEA). Continuous electroencephalography, monitored by neurophysiological experts, is used to detect cerebral ischemia during surgery; however, real-time visual interpretation is resource-intensive and error-prone. We evaluated machine learning (ML) models, including random forest (RF), eXtreme Gradient Boosting with a random forest base classifier (XGB), elastic-net logistic regression (LR), support vector classifier (SVC) with a radial basis function kernel, and naive Bayes (NB) classifier, for automated detection of cerebral ischemia during CEA using quantitative electroencephalographic (qEEG) features. RF achieved the highest sensitivity (0.79–0.83) and an area under the precision–recall curve (AUPRC) of 0.44, while XGB demonstrated the highest specificity (0.93–0.96) with an AUPRC of 0.36. Both models showed high negative predictive values and high area under the receiver operating characteristic (AUROC) scores. Feature-importance analysis identified alpha-band activity and hemispheric asymmetry as the most discriminative qEEG predictors of ischemia. These results highlight the potential of ML-assisted monitoring to support neurophysiology experts and enhance patient safety during high-risk surgical procedures.

## Introduction

Over the past decade, the risk of perioperative stroke, occurring during surgery or within 30 days afterward, has increased, despite advances in surgical techniques and monitoring during surgery^1^. Perioperative strokes affect approximately 50,000 patients in the United States (U.S.) each year and are associated with high morbidity and mortality, often resulting in long-term declines in quality of life. Most strokes occur during the surgery itself, with rates up to 6% for cardiovascular and neurological surgeries, and around 1% for general surgical procedures^2, 3^. Cardiovascular and neurological surgeries that are at high-risk account for roughly one million of the 50 million surgical procedures performed annually in the U.S. Reducing the risk of perioperative strokes, especially in high-risk surgeries, would not only enhance patient safety but also help alleviate significant healthcare costs^1, 2^.

During surgery, maintaining uninterrupted cerebral blood flow (CBF) is essential for keeping the brain functioning by supplying oxygen and nutrients. The brain has very limited reserves, so even short interruptions in CBF can lead to serious injury. When CBF is temporarily reduced or stopped, it causes ischemia, which can lead to reversible brain injury. However, if the disruption in CBF is prolonged, it causes stroke, which often results in irreversible brain damage. The progression from ischemia to stroke can occur within minutes or take several hours, depending on the severity and duration of the CBF reduction^4, 5^. Therefore, detection of cerebral ischemia is crucial to allow for timely interventions that can prevent strokes.

Continuous electroencephalography (cEEG) is a non-invasive method for intraoperative neurophysiological monitoring (IONM) of the brain. It continuously measures and records cerebral electrical activity by detecting voltage changes over time via electrodes placed at standardized scalp locations^6, 7^. The electrodes are labeled with a letter and a number: the letter indicates the underlying cerebral region (F for frontal, P for parietal, T for temporal, and O for occipital), and even numbers correspond to the right hemisphere and odd numbers to the left hemisphere. For example, F3 is located over the left frontal lobe, and O2 is situated over the right occipital lobe. The voltage measured by cEEG is recorded between pairs of electrodes, with each pair creating a "channel." For instance, the F3–P3 channel reflects the voltage difference between the electrodes positioned over the left frontal and left parietal lobes (Figure 1).

**Figure 1.**
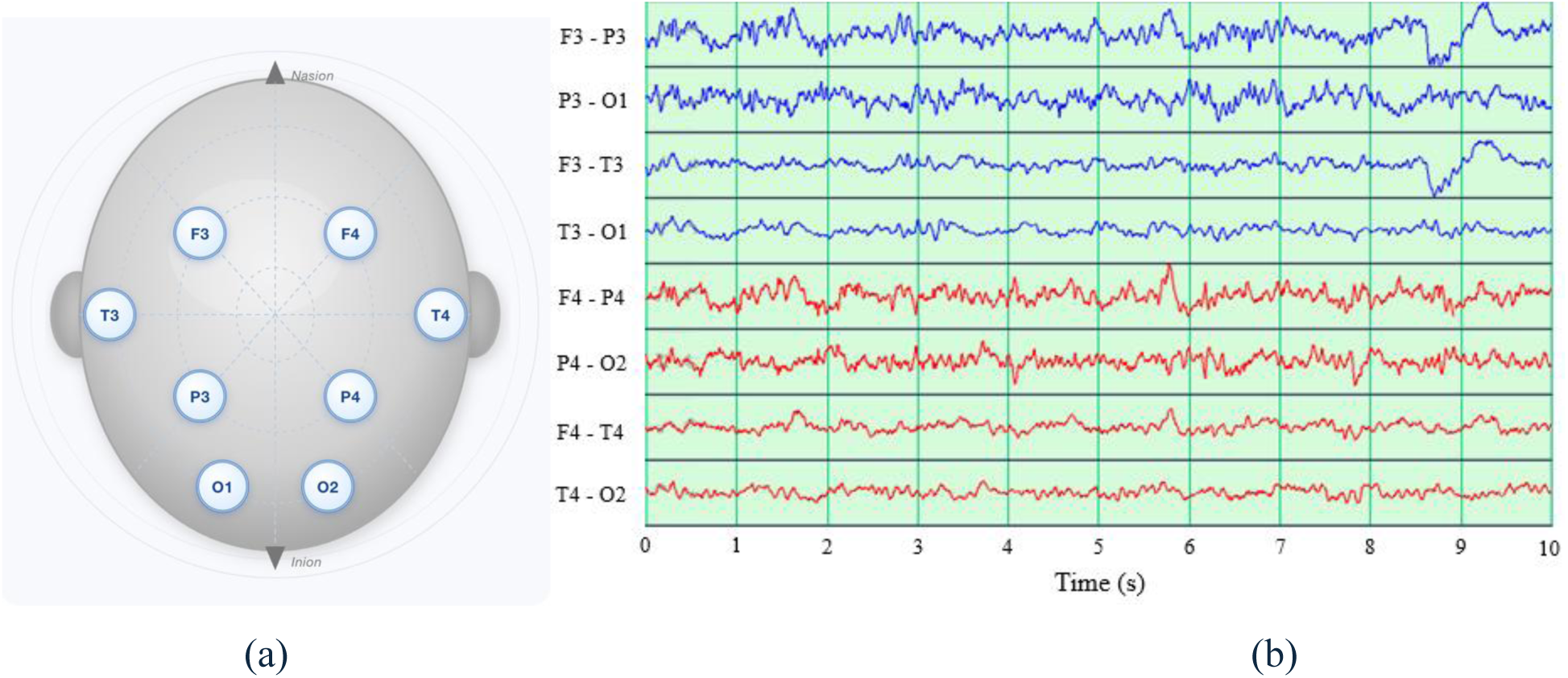
(a) Scalp locations for electrodes F3, P3, T3, O1, F4, P4, T4, and O2. b) An eight-channel continuous EEG segment is shown. Channels F3–P3, P3–O1, F3–T3, and T3–O1 (blue) measure electrical activity from the left hemisphere, while channels F4–P4, P4–O2, F4–T4, and T4–O2 (red) measure electrical activity from the right hemisphere.

For the past few decades, IONM using cEEG has been employed by anesthesiologists to assess the depth of anesthesia and by neurophysiologists to monitor cerebral ischemia^3, 8^. This was made possible by technological improvements in the 1990s, which enabled the digitization and electronic storage of cEEG signals, allowing for continuous monitoring over extended periods. In high-risk surgical procedures that may disrupt CBF, such as carotid endarterectomy (CEA), cEEG is now routinely utilized to detect ischemia^3^. CEA is performed to remove atherosclerotic plaque from the common or internal carotid artery and requires temporary carotid artery clamping. During and immediately after clamping, the brain is especially vulnerable to decreased CBF and ischemic injury. Beyond CEA, intraoperative cEEG is increasingly used in other high-risk cardiovascular and neurosurgical procedures where cerebral ischemia is a concern.

Intraoperative monitoring personnel, such as neurophysiologists, visually interpret cEEG waveforms by evaluating changes in amplitude and morphology across specific frequency bands. The standard frequency bands are as follows: delta (0–4 Hz), theta (4–8 Hz), alpha (8–12 Hz), beta (12–30 Hz), and gamma (30–100 Hz), listed in order of increasing frequency. Generally, as frequency increases, cEEG amplitude decreases. The delta and theta bands have low frequencies and larger amplitudes, often referred to as "slow activity," while the alpha, beta, and gamma bands have higher frequencies and smaller amplitudes, known as "fast activity"^9^.

Real-time visual interpretation of cEEG by neurophysiologists is resource-intensive, requires substantial expertise, and is prone to error^10^. These challenges limit the scalability of cEEG monitoring in the operating room. Computer-assisted detection using machine learning (ML) has the potential to rapidly and accurately identify ischemia, augmenting human monitoring.

To quantify cEEG patterns indicative of ischemia, particularly in CEA, several quantitative metrics have been developed to characterize brain activity. These quantitative electroencephalographic metrics are derived from digitally recorded cEEG signals to provide objective, reproducible measures of brain activity. In the context of cerebral ischemia, qEEG parameters have been established to detect subtle, often transient changes in cerebral activity that may be overlooked through visual inspection alone. By applying computational techniques such as the fast Fourier transform (FFT), cEEG signals can be decomposed into their constituent frequency components, enabling quantification of power within specific frequency bands^11^.

The qEEG metrics are divided into two main categories: frequency-based and amplitude-based. Frequency-based qEEG metrics are usually derived from the continuous EEG (cEEG) power spectrum using Fast Fourier Transform (FFT). The power within a specific frequency band, for instance, the alpha band (8–12 Hz), is determined by summing the spectral power across that frequency range. Informative ratios of high-to-low-frequency power include the alpha-to-delta ratio (ADR), beta-to-delta ratio (BDR), and alpha-beta-to-delta-theta ratio (ABDTR)^12^. Another commonly used metric is the spectral edge frequency (SEF), defined as the frequency below which a specified proportion (typically 90%) of the total power is contained^13^. Because ischemia and stroke often affect only one hemisphere, asymmetry metrics such as the pairwise-derived brain symmetry index (pd-BSI) have been developed. Among amplitude-based metrics, the most widely used is amplitude-integrated EEG (aEEG), which represents the amplitude of filtered and smoothed EEG signals^14^. Table 1 summarizes key qEEG parameters relevant to ischemia detection and outlines how these measures are affected by cerebral ischemia.

**Table 1.** Selected qEEG parameters, brief description, and reported effect in cerebral ischemia.

| qEEG parameter | Description | Effect in ischemia |
| --- | --- | --- |
| Power (band-specific) | Power within a defined EEG frequency band (delta, theta, alpha, beta, gamma). | Increase in low-frequency power (delta, theta) and reduction in higher-frequency power (alpha, beta, gamma), reflecting ischemia-induced cortical slowing. |
| Power ratios | Ratios comparing higher-frequency to lower-frequency band power (e.g., ADR, BDR, ABDTR). | Ratios emphasizing higher frequencies typically decrease during ischemia due to the dominance of slow-wave activity |
| Spectral edge frequency (SEF90) | Frequency below which 90% of the total EEG power is contained. | SEF90 decreases during ischemia, indicating a shift of spectral power toward lower frequencies. |
| Pairwise-derived brain symmetry index (pd-BSI) | Interhemispheric asymmetry computed from power differences between homologous electrode pairs. | Focal ischemia increases hemispheric asymmetry, leading to elevated pd-BSI values. |
| Amplitude-integrated EEG (aEEG) | Amplitude range of EEG activity displayed as a time-compressed trend. | Reduced EEG amplitude and narrowing of the aEEG band are commonly observed during ischemia. |

Ischemic changes during CEA have been well documented in numerous studies. In one study involving 92 CEA patients, Visser et al. utilized cEEG monitoring combined with transcranial Doppler ultrasonography to assess intracranial CBF^15^. They found that after carotid clamping, severe ischemia was indicated by reductions in alpha- and beta-band activity, accompanied by increases in delta- and theta-band activity. In contrast, mild ischemia was associated only with a decrease in alpha activity. In some patients, alpha and beta activity increased while delta and theta activity decreased, which is attributed to arousal due to pain or hemodynamic stress induced by the clamping. Kamitaki et al., analyzing cEEG from 118 CEA patients, found that reductions in alpha, beta, and theta power of 52.1%, 41.6%, and 36.4%, respectively, were the strongest predictors of ischemia after clamping^11^. In a large study of 1,551 patients, Pedapati et al. evaluated 10 qEEG metrics and observed significant decreases in alpha, beta, theta, delta, and gamma power, as well as ADR, BDR, ABDTR, SEF90, and aEEG when ischemia was evident on visual EEG inspection^16^.

## Results

In this section, we describe the data characteristics, feature selection, and report model performance with both the full and a reduced set of features.

### Description of data and features

We used a large retrospective dataset of patients who underwent CEA with cEEG monitoring at a major academic healthcare system that comprises multiple hospitals. The full dataset comprised recordings from 1,611 patients who underwent CEA between 2009 and 2019. For each patient in the dataset, a 10-minute cEEG segment beginning at the time of carotid clamping was extracted. Each segment was divided into non-overlapping 20-second intervals, from which 111 qEEG-based features were extracted.

Feature extraction proceeded as follows. For each of the eight channels, we computed six band powers (delta, theta, alpha, beta, gamma, wideband), three power ratios (ADR, BDR, ABDTR), the spectral edge frequency at 90% power, and the aEEG (Table 2). In addition, we calculated 10 averaged features from the four left-hemisphere channels, 10 from the four right-hemisphere channels, and 10 from averaging over both right and left (Table 2). A final feature, the pairwise-derived brain symmetry index (pd-BSI), was computed from power values across all channels and frequency bands.

**Table 2.**
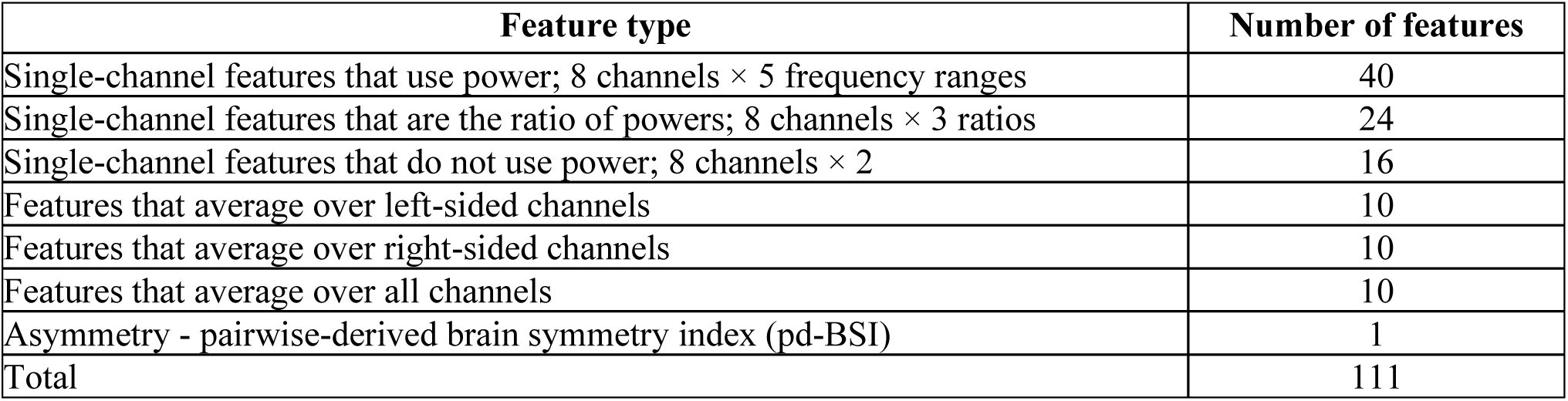
Summary of 111 qEEG features computed from each 20-second interval.

| Feature type | Number of features |
| --- | --- |
| Single-channel features that use power; 8 channels × 5 frequency ranges | 40 |
| Single-channel features that are the ratio of powers; 8 channels × 3 ratios | 24 |
| Single-channel features that do not use power; 8 channels × 2 | 16 |
| Features that average over left-sided channels | 10 |
| Features that average over right-sided channels | 10 |
| Features that average over all channels | 10 |
| Asymmetry - pairwise-derived brain symmetry index (pd-BSI) | 1 |
| Total | 111 |

The dataset was partitioned into training and test sets. The training set comprised 24,090 intervals from 803 patients, of whom 48 (2.4% of intervals) had confirmed ischemic events. The test set comprised 24,240 intervals from 808 patients, of whom 40 (1.8% of intervals) had confirmed ischemic events.

### Feature selection

Figure 2 presents the top 20 permutation-based feature importance scores computed on the validation set, ranked by mean decrease in model performance following random permutation. Features exhibiting positive importance values demonstrated a measurable contribution to predictive performance, while those with near-zero or negative values were excluded from further analysis. The 65 retained features were those most consistently associated with model performance within this evaluation framework (see Table S2 in Supplementary Information). Although individual importance values were modest in magnitude, this pattern reflects the distributed nature of EEG-derived predictors rather than the dominance of any single feature; importance scores were therefore interpreted in terms of relative ranking and consistency across permutations.

**Figure 2.**
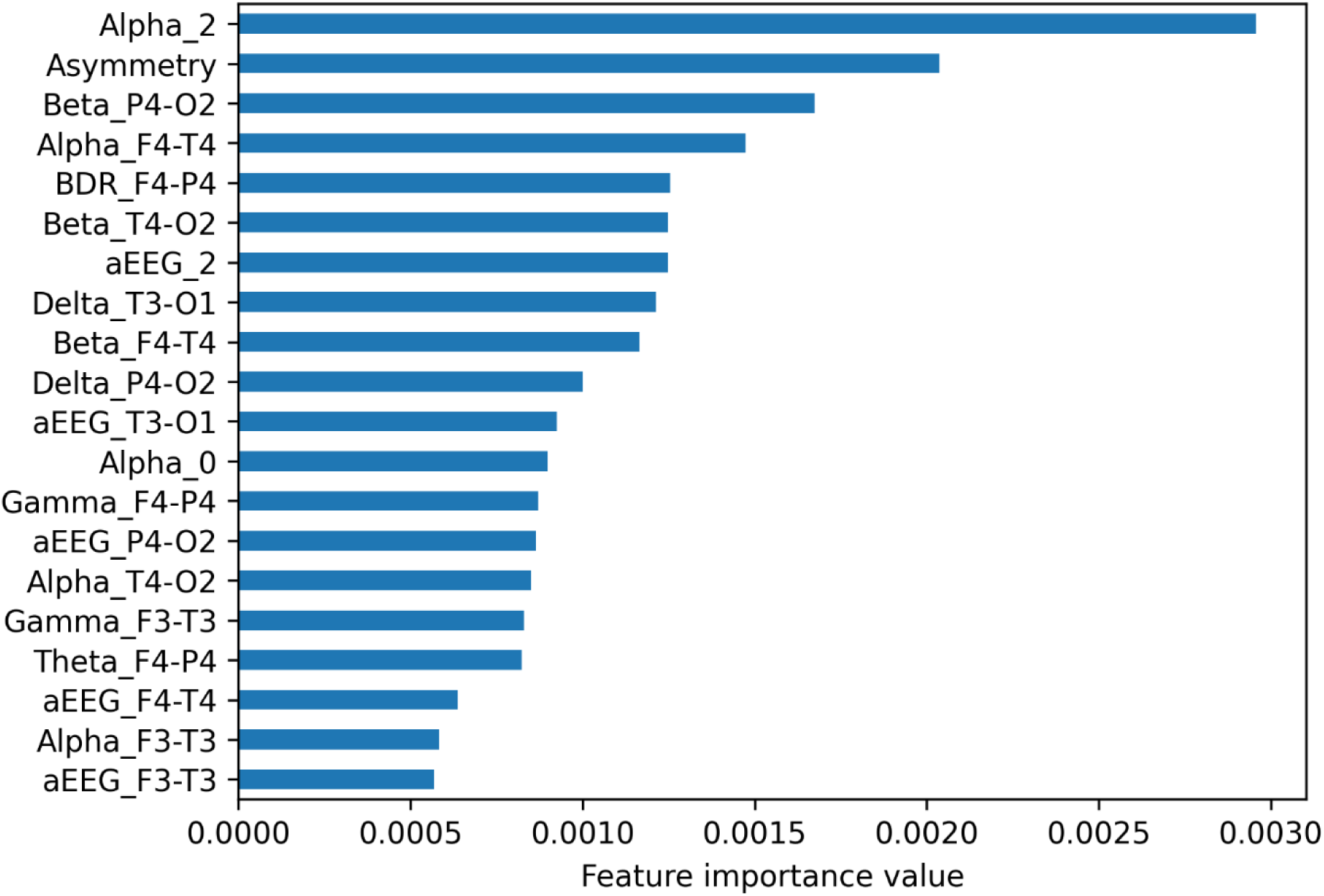
Top 20 qEEG features ranked by importance score. Alpha_2 has the highest score, followed by Asymmetry and Beta_P4–O2. Mid-ranked features include channel-specific Alpha, Beta, and Delta band power from right hemisphere electrodes (F4, T4, P4, and O2). Features derived from aEEG and those in the Gamma frequency band appear near the lower end of the ranking.

As a complementary dimensionality reduction strategy, we applied principal component analysis (PCA). Model performance was evaluated across a predefined range of component counts (5, 10, 15, 20, 30, 40, 50, and 60), with the optimal number selected independently for each model based on predictive performance. This approach enabled systematic assessment of the trade-off between dimensionality reduction and classification accuracy.

### Predictive performance

We evaluated several supervised ML algorithms to develop classification models, including random forest^17^ (RF), eXtreme Gradient Boosting with a random forest^18^ base classifier (XGB), elastic-net logistic regression^19^ (LR), support vector classifier (SVC) with a radial basis function kernel^20^, and naive Bayes^20^ (NB) classifier. Model performance was assessed across three experimental settings: all 111 features, a reduced set of selected features, and features derived from principal component analysis. Model performance was evaluated across three experimental configurations: a baseline using the full 111-feature set, a reduced configuration retaining only selected features, and a dimensionality-reduced configuration based on principal component analysis (PCA). Across all settings, threshold-dependent metrics (sensitivity, specificity, PPV, and NPV) were computed at the optimal operating threshold determined by the Youden J index^21^, while threshold-free performance was characterized by AUROC and AUPRC. Complete results, including 95% confidence intervals, are reported in Tables 3, 4, and 5.

**Table 3.** Model performance using all qEEG features. Values in parentheses represent 95% confidence intervals. Preference metrics include AUROC = area under the receiver operating characteristic curve; AUPRC = area under the precision-recall curve; Sens. = sensitivity; Spec. = specificity. = specificity; Prec./PPV = precision / positive predictive value; NPV = negative predictive value. Models include RF = random forest; XGB = Extreme Gradient Boosting, LR = logistic regression, SVC = support vector classifier, NB = naïve Bayes.

| Models | AUROC | AUPRC | Sens. | Spec. | Prec./PPV | NPV |
| --- | --- | --- | --- | --- | --- | --- |
| RF | **0.93 (0.92–0.95)** | **0.42 (0.36–0.46)** | **0.83 (0.79–0.87)** | 0.93 (0.92–0.93) | 0.17 (0.15–0.18) | **0.997 (0.996–0.997)** |
| XGB | 0.92 (0.91–0.94) | 0.35 (0.31–0.40) | 0.78 (0.74–0.81) | **0.94 (0.94–0.95)** | **0.20 (0.18–0.22)** | 0.996 (0.995–0.997) |
| LR | 0.86 (0.84–0.88) | 0.17 (0.14–0.20) | 0.65 (0.60–0.69) | 0.92 (0.92–0.93) | 0.13 (0.12–0.15) | 0.993 (0.992–0.994) |
| SVC | 0.87 (0.85–0.89) | 0.24 (0.20–0.28) | 0.68 (0.63–0.72) | 0.92 (0.92–0.93) | 0.14 (0.12–0.15) | 0.994 (0.993–0.995) |
| NB | 0.83 (0.80–0.85) | 0.11 (0.09–0.12) | 0.75 (0.70–0.79) | 0.82 (0.81–0.82) | 0.07 (0.06–0.08) | 0.995 (0.993–0.996) |

**Table 4.** Model performance using only qEEG features identified through feature selection. Values in parentheses represent 95% confidence intervals.

| Models | AUROC | AUPRC | Sens. | Spec. | Prec./PPV | NPV |
| --- | --- | --- | --- | --- | --- | --- |
| RF | **0.93 (0.91–0.94)** | **0.40 (0.35–0.45)** | **0.79 (0.75–0.83)** | 0.94 (0.93–0.94) | 0.18 (0.17–0.20) | **0.996 (0.995–0.997)** |
| XGB | 0.91 (0.90–0.93) | 0.29 (0.25–0.33) | 0.69 (0.64–0.74) | **0.96 (0.96–0.96)** | **0.23 (0.21–0.26)** | 0.994 (0.993–0.995) |
| LR | 0.87 (0.85–0.89) | 0.17 (0.14–0.20) | 0.60 (0.54–0.65) | 0.93 (0.93–0.94) | 0.14 (0.13–0.16) | 0.992 (0.991–0.993) |
| SVC | 0.86 (0.84–0.88) | 0.22 (0.19–0.26) | 0.72 (0.67–0.76) | 0.90 (0.89–0.90) | 0.11 (0.10–0.12) | 0.994 (0.993–0.995) |
| NB | 0.83 (0.80–0.86) | 0.14 (0.12–0.16) | 0.76 (0.72–0.80) | 0.81 (0.81–0.82) | 0.07 (0.06–0.07) | 0.995 (0.994–0.996) |

**Table 5.** Model using principal components derived through application of principal component analysis (PCA) to all qEEG features. Values in parentheses represent 95% confidence intervals.

| Models | AUROC | AUPRC | Sens. | Spec. | Prec./PPV | NPV |
| --- | --- | --- | --- | --- | --- | --- |
| RF | **0.92 (0.91–0.94)** | **0.44 (0.39–0.49)** | **0.82 (0.78–0.85)** | 0.91 (0.90–0.91) | 0.14 (0.12–0.15) | **0.996 (0.996–0.997)** |
| XGB | 0.89 (0.87–0.91) | 0.36 (0.32–0.41) | 0.68 (0.64–0.72) | **0.94 (0.93–0.94)** | **0.16 (0.14–0.17)** | 0.994 (0.993–0.995) |
| LR | 0.86 (0.84–0.88) | 0.15 (0.13–0.18) | 0.69 (0.64–0.73) | 0.89 (0.89–0.90) | 0.10 (0.09–0.11) | 0.994 (0.993–0.995) |
| SVC | 0.89 (0.87–0.92) | 0.41 (0.36–0.46) | 0.77 (0.73–0.81) | 0.93 (0.92–0.93) | 0.16 (0.14–0.17) | 0.996 (0.995–0.996) |
| NB | 0.87 (0.86–0.89) | 0.13 (0.11–0.15) | 0.67 (0.63–0.72) | 0.89 (0.89–0.90) | 0.10 (0.09–0.11) | 0.993 (0.992–0.995) |

Using the full feature set (Table 3), RF and XGB achieved the highest AUROC, sensitivity, and PPV among the five classifiers evaluated. RF achieved the highest AUPRC, a difference that was statistically significant compared with all other models, including XGB. Given the low prevalence of positive cases in the dataset, all classifiers exhibited high specificity and NPV, reflecting the class imbalance rather than differential discriminative ability.

Results using the top 65 permutation-selected features were broadly comparable to those obtained with the full feature set. RF and XGB again ranked as the best-performing models; however, XGB surpassed RF on specificity and PPV under this configuration. As with the full-feature results, RF achieved the highest AUPRC, with a statistically significant margin over all other models, including XGB.

PCA-based models (Table 4) were evaluated across component counts of 5, 10, 15, 20, 30, 40, 50, 60, 70, 80, 90, and 100, with the optimal number selected independently for each classifier. Overall performance was again comparable to the full-feature baseline. RF led on AUROC, AUPRC, and sensitivity, while XGB and SVC achieved the highest PPV under this configuration.

Across all three experimental settings, feature selection and PCA produced modest and variable effects on both discriminative and classification metrics, with no configuration yielding uniformly superior performance across all evaluated criteria.

## Discussion

The results demonstrate the potential of ML to detect cerebral ischemia during CEA using qEEG features, addressing a critical need for automated, real-time IONM. Our findings provide several important insights into model performance, feature importance, and the clinical applicability of computer-assisted ischemia detection.

Among the ML models using all features, RF and XGB achieved the highest AUPRCs of 0.44 and 0.36, respectively. These results indicate that ensemble tree models are particularly effective at capturing complex, nonlinear relationships among qEEG parameters and ischemic events. When the features were reduced to 65, RF’s performance decreased slightly to an AUPRC of 0.40, while XGB’s performance decreased moderately to 0.29. This indicates that some information was lost when reducing the features from 111 to 65. Interestingly, when PCA was applied, the performance of both RF and XGB improved slightly, reaching AUPRC values of 0.44 and 0.36, respectively. PCA enhances classifier performance by removing redundant and noisy features and transforming correlated features into a smaller set of uncorrelated components that capture most of the data’s variation. This process can help reduce overfitting and improve classifier performance.

Across all three feature representations, RF achieved the highest sensitivity, ranging from 0.79 to 0.83. In contrast, XGB achieved the highest specificity, ranging from 0.93 to 0.96. Overall, PPV is low across all models, with XGB achieving the highest PPV (0.16-0.23) across all three feature representations. The consistently low PPV across all models reflects the inherent difficulty of detecting rare events in a highly imbalanced dataset, where ischemic intervals account for only about 2% of all intervals. In the context of CEA, however, a lower PPV is clinically acceptable because the primary objective is to avoid missing patients who are at high risk of a stroke when the carotid artery is clamped. Moreover, the corrective response, releasing the clamp, inserting a shunt, and continuing the procedure, adds a limited procedural burden.

In a class-imbalanced setting, the overall performance of RF and XGB is clinically useful. The models identify a substantial proportion of true ischemic events while maintaining a manageable false-positive rate in practice. The results underscore the challenge of detecting subtle cEEG changes indicative of early ischemia before they progress to more pronounced cEEG abnormalities suggestive of stroke.

Overall, the NPV is consistently high across all models and feature representations, demonstrating a strong ability to rule out ischemia when the model does not predict it. The high NPV addresses a major limitation of current visual cEEG interpretation: the resource-intensive nature of continuous monitoring and the risk of human error during extended surgical procedures. Our findings support the potential for these ML models to augment, rather than replace, the interpretation by neurophysiologists. The automated system could serve as a sensitive first-line screening tool, alerting neurophysiologists to potential ischemic changes that warrant further investigation. This approach would allow neurophysiologists to focus their expertise on ambiguous cases and decision-making regarding interventions, while the model continuously monitors cEEG signals.

The feature importance analysis shows that alpha band activity is the most significant indicator for detecting ischemia. This aligns with established neurophysiological knowledge, as previous studies^15^ have documented that mild ischemia during CEA is associated with reductions in alpha activity. The significance of alpha band features suggests that changes in this frequency range may be among the earliest and most sensitive markers of reduced CBF. Asymmetry measures ranked second among the important features, supporting the clinical observation that ischemia during CEA often affects one hemisphere more than the other. The development and validation of asymmetry metrics, such as the pd-BSI, reflect the hemispheric nature of carotid artery disease and the localized impact of temporary clamping. This lateralization provides an internal control, as the contralateral hemisphere continues to receive adequate perfusion, making asymmetry a robust indicator of unilateral ischemic changes.

Beta band activity recorded from multiple electrode locations also emerged as an important indicator for detecting ischemia, aligning with previous research indicating that reductions in beta power are strong predictors of post-clamp ischemia. The involvement of both frontal and posterior beta activity suggests that ischemia impacts multiple cortical regions. This may reflect varying vulnerability patterns or the gradual spread of ischemic changes over time.

Several limitations warrant consideration when interpreting these findings. First, the study was based on data from a limited number of surgical centers, which may affect generalizability to different patient populations, surgical techniques, and cEEG recording protocols. Validation on external, multicenter datasets would strengthen confidence in the models’ real-world performance and identify any institution-specific biases in the training data. Second, the temporal dynamics of ischemia development were not explicitly modeled in this analysis. Ischemic changes often evolve gradually, and incorporating time-series modeling might improve early detection by identifying trends and trajectories in qEEG parameters. Future work should explore whether temporal features, such as rate of change in power spectral density or evolving asymmetry patterns, enhance predictive performance. Third, the class imbalance indicates that ischemic events account for only a small proportion of the total monitoring time. While our models achieved excellent AUROC, the lower AUPRC and PPV suggest opportunities for improvement through advanced techniques such as synthetic minority oversampling, cost-sensitive learning, or ensemble methods specifically designed for imbalanced datasets. Optimizing the decision threshold based on clinical risk tolerance could also improve the balance between sensitivity and specificity. The ground truth for ischemia in this study was based on annotations made by the monitoring neurophysiologist in the operating room. However, this may not be optimal, as the neurophysiologist is under significant time pressure during surgery. A more accurate gold standard could be established by having experts review the cEEG post-operatively, when they are not constrained by time.

This study demonstrates that ML models, particularly RF and XGB, can accurately detect cerebral ischemia during CEA using cEEG. Alpha band activity and hemispheric asymmetry emerged as the most discriminative predictors, consistent with the established understanding of ischemic cEEG changes. The high AUROC and NPV support the potential clinical utility of automated ischemia-detection systems in enhancing patient safety during high-risk surgeries. While challenges remain in optimizing sensitivity and addressing class imbalance, these findings represent a meaningful step toward scalable, real-time computer-assisted neuromonitoring that could reduce the burden on specialized personnel and improve outcomes for surgical patients at risk of perioperative stroke.

## Methods

In this section, we describe the dataset and the feature engineering used for the study, how labels were obtained, the ML methods, the experimental design, and the evaluation.

### Data and feature engineering

The dataset comprised cEEG recordings from 1,611 patients who underwent CEA between 2009 and 2019. Signals were acquired at a sampling rate of 500 Hz across eight bipolar channels: F3– P3, P3–O1, F3–T3, and T3–O1 from the left hemisphere, and F4–P4, P4–O2, F4–T4, and T4–O2 from the right hemisphere.

Raw signals were preprocessed using a standard filter cascade comprising a high-pass filter at 0.1 Hz, a low-pass filter at 70 Hz, and a 60-Hz notch filter to attenuate line noise. For each patient, a 10-minute cEEG segment beginning at the moment of carotid clamping was then extracted and divided into non-overlapping 20-second intervals. A total of 111 qEEG-derived features were computed per interval. Frequency-domain features were obtained from the power spectrogram, calculated by applying a short-time Fourier transform (FFT) to a sliding window; spectral power values were subsequently converted to decibels using the transformation 10 × ln(power). Full feature definitions are provided in Table S1 of the Supplementary Information.

All features were normalized at the patient level relative to a 120-second pre-clamp baseline. For each patient, candidate baseline segments, defined as all 120-second intervals ending immediately before clamp onset, were evaluated, and the segment exhibiting minimal noise and variance was selected. Baseline distribution parameters estimated from this segment were used to compute per-feature means and standard deviations, and all feature values were subsequently transformed into z-scores. Each 20-second interval was assigned a binary label: positive (ischemic) if more than half its duration overlapped with a neurophysiologist-annotated ischemic period, and negative (non-ischemic) otherwise.

### Machine learning methods

We developed and evaluated several supervised ML algorithms, including RF, XGB, LR, SVC, and NB. Our primary focus was on tree-based methods, specifically RF and gradient-boosting approaches such as XGB, given evidence that these models outperform alternatives, including deep learning, on tabular data. Although direct comparisons between tree-based and deep learning models for EEG-derived features remain limited, available evidence suggests that tree-based models can achieve competitive or superior performance in this domain, as demonstrated in studies predicting drowsiness from EEG signals.

To provide a robust automated benchmark, we additionally evaluated AutoGluon^22^, an open-source automated machine learning (AutoML) framework optimized for tabular prediction. AutoGluon automates model selection, hyperparameter tuning, and ensemble construction by training a diverse set of base classifiers—spanning tree-based, linear, and neural-network architectures—and combining them into multi-layer stacked ensembles. Its capacity to deliver strong performance on structured datasets with minimal manual configuration makes it a compelling reference point for assessing the ceiling of automated supervised learning in this setting (see Table S3 in Supplementary Information for results).

For all models, predicted probabilities were dichotomized using the threshold on the receiver operating characteristic curve that maximized the Youden J index, defined as sensitivity + specificity − 1. This criterion identifies the point of greatest improvement over random discrimination, geometrically, the maximum vertical distance from the receiver operating characteristic (ROC) curve to the no-discrimination diagonal, and implicitly assumes equal costs for false positives and false negatives.

### Feature selection and dimensionality reduction

In addition to using all 111 features, we evaluated classifiers with a reduced feature set. To identify which features significantly contributed to model performance, we first employed permutation importance. This method assesses the relevance of each feature by measuring the decrease in model accuracy when the values of that feature are randomly shuffled, thereby disrupting its relationship with the target variable^17^. Second, we applied PCA, which transforms the original features into a smaller set of uncorrelated principal components that capture most of the data’s variance^23^.

### Evaluation methods

We assessed the performance of the models using both threshold-based metrics. Threshold-dependent metrics, including sensitivity, specificity, PPV, and NPV, were computed at the optimal operating threshold for each model, determined via the Youden J index. Threshold-free performance was characterized by AUROC and AUPRC.

AUROC is the most used threshold-free metric for evaluating ML models, as it measures a model’s ability to distinguish between positive and negative samples across various probability thresholds and is not influenced by the target distribution. However, AUPRC has gained importance in scenarios with imbalanced datasets like ours, as it effectively balances precision and recall and is sensitive to the target distribution. In particular, AUPRC is often considered more informative than AUROC in rare-event settings, where there may be few positive targets. In these settings, the ROC curve can present a misleadingly optimistic view of model performance, whereas the precision-recall (PR) curve offers a more accurate assessment.

### Experimental setup

The ML models were developed from the training set using 80/10/10 splits for training/validation/test. All models were calibrated using isotonic scaling. The ML models were then evaluated on the test dataset. We report performance on the test dataset. Data preprocessing was implemented in Python 3.12.7, and model development was performed using scikit-learn version 1.4.2.

## Ethics approval

The Institutional Review Board at the University of Pittsburgh approved this study (STUDY25100049 and STUDY25100194).

## Data availability

The data generated and analyzed in the current study are not publicly available due to patient privacy regulations but are available from the corresponding authors upon reasonable request.

## Code availability

The underlying code for this study is not publicly available but may be made available to qualified researchers from the corresponding authors on reasonable request.

## Acknowledgements

The research reported in this publication was supported by the National Center for Advancing Translational Sciences of the National Institutes of Health under award number UL1 TR001857. The content is solely the responsibility of the authors and does not necessarily represent the official views of the National Institutes of Health.

## Author contributions

S.V. and M.N. led the conceptualization of the study and wrote the initial draft of the manuscript. A.I.M. and J.U.E. assisted with preprocessing and feature engineering. K.B. assisted with the analyses. P.D.T. collected and verified the raw data and provided the annotations. All authors had full access to the data and shared final responsibility for the decision to submit the manuscript for publication.

## References

1 Shu, L. et al. Perioperative stroke: Mechanisms, risk stratification, and management. Stroke (2025).

2. Thirumala, P. D., Thiagarajan, K., Gedela, S., Crammond, D. J. & Balzer, J. R. Diagnostic accuracy of eeg changes during carotid endarterectomy in predicting perioperative strokes. J. Clin. Neurosci. 25, 1–9 (2016).

3. Zhou, Z.-B., Meng, L., Gelb, A. W., Lee, R. & Huang, W.-Q. Cerebral ischemia during surgery: an overview. J. Biomed. Res. 30, 83 (2016).

4. Sacco, R. L. et al. An updated definition of stroke for the 21st century: a statement for healthcare professionals from the American Heart Association/American Stroke Association. Stroke 44, 2064–2089 (2013).

5. Sharbrough, F. W., Messick Jr, J. M. & Sundt Jr, T. M. Correlation of continuous electroencephalograms with cerebral blood flow measurements during carotid endarterectomy. Stroke 4, 674–683 (1973).

6. Hossmann, K.-A. Viability thresholds and the penumbra of focal ischemia. Annals Neurol. Off. J. Am. Neurol. Assoc. Child Neurol. Soc. 36, 557–565 (1994).

7. Sun, Y., Wei, C., Cui, V., Xiu, M. & Wu, A. Electroencephalography: clinical applications during the perioperative period. Front. medicine 7, 251 (2020).

8. Chiappa, K. H., Burke, S. R. & Young, R. R. Results of electroencephalographic monitoring during 367 carotid endarterectomies. use of a dedicated minicomputer. Stroke 10, 381– 388 (1979).

9. Sanei, S. & Chambers, J. A. EEG signal processing (John Wiley & Sons, 2013).

10. Zhao, G. et al. Application of quantitative electroencephalography in predicting early cerebral ischemia in patients undergoing carotid endarterectomy. Front. Neurol. 14, 1159788 (2023).

11. Kamitaki, B. K., Tu, B., Wong, S., Mendiratta, A., & Choi, H. Quantitative EEG changes correlate with post-clamp ischemia during carotid endarterectomy. J. Clin. Neurophysiol. 38, 213–220 (2021).

12. Rots, M., Van Putten, M., Hoedemaekers, C. & Horn, J. Continuous EEG monitoring for early detection of delayed cerebral ischemia in subarachnoid hemorrhage: a pilot study. Neurocritical care 24, 207–216 (2016).

13. McKeever, S., Johnston, L. & Davidson, A. An observational study exploring amplitude-integrated electroencephalogram and spectral edge frequency during paediatric anaesthesia. Anaesth. Intensive Care 40, 275–284 (2012).

14. Maynard, D., Prior, P. & Scott, D. A continuous monitoring device for cerebral activity. Electroencephalogr. Clinical Neurophysiology 27, 672–673 (1969).

15. Visser, G., Wieneke, G. & Van Huffelen, A. Carotid endarterectomy monitoring: patterns of spectral EEG changes due to carotid artery clamping. Clin. Neurophysiology 110, 286– 294 (1999).

16. Pedapati, V. et al. Quantitative EEG changes in carotid endarterectomy correlated with ischemia. In 2022 IEEE Signal Processing in Medicine and Biology Symposium (SPMB), 1–5 (IEEE, 2022).

17. Breiman, L. Random forests. Mach. learning 45, 5–32 (2001).

18. Chen, T. XGBoost: A scalable tree boosting system. Cornell Univ. (2016).

19. Hosmer Jr, D. W., Lemeshow, S. & Sturdivant, R. X. Applied logistic regression (John Wiley & Sons, 2013).

20. Hastie, T., Tibshirani, R., Friedman, J., et al. The elements of statistical learning (2009).

21. Šimundić, A.-M. Measures of diagnostic accuracy: basic definitions. ejifcc 19, 203 (2009).

22. Erickson, N., et al. Autogluon-tabular: Robust and accurate automl for structured data. arXiv preprint arXiv:2003.06505 (2020).

23. Jolliffe, I. T. & Cadima, J. Principal component analysis: a review and recent developments. Philos. Trans. R. Soc. A: Math. Phys. Eng. Sci. 374, 20150202 (2016).

