## Supplementary information for "Machine learning to detect intraoperative ischemia from electroencephalography in carotid endarterectomy surgery"

**Table S1.** List of 111 qEEG features computed from each 20-second interval. The frequency bands are defined as follows: delta (0–4 Hz), theta (4–8 Hz), alpha (8–12 Hz), beta (12–30 Hz), and gamma (30–100 Hz). Channels F3–P3, P3–O1, F3–T3, and T3–O1 measure electrical activity from the left hemisphere, while channels F4–P4, P4–O2, F4–T4, and T4–O2 measure electrical activity from the right hemisphere. The qEEG feature naming follows a standardized convention. For example, Delta_T3–O1 denotes the power in the delta frequency band (0–4 Hz) measured from the T3–O1 channel on the left side of the head during a 20-second interval.

| Feature type | Number of features | Description of features |
|---|---|---|
| Single-channel features that use power | 8 channels × 5 frequency ranges = 40 | Delta_T3–O1<br>Delta_P3–O1<br>Delta_F3–T3<br>Delta_F3–P3<br>Delta_F4–P4<br>Delta_F4–T4<br>Delta_P4–O2<br>Delta_T4–O2<br><br>Theta_T3–O1<br>Theta_P3–O1<br>Theta_F3–T3<br>Theta_F3–P3<br>Theta_F4–P4<br>Theta_F4–T4<br>Theta_P4–O2<br>Theta_T4–O2<br><br>Alpha_T3–O1<br>Alpha_P3–O1<br>Alpha_F3–T3<br>Alpha_F3–P3<br>Alpha_F4–P4<br>Alpha_F4–T4<br>Alpha_P4–O2<br>Alpha_T4–O2<br><br>Beta_T3–O1<br>Beta_P3–O1<br>Beta_F3–T3<br>Beta_F3–P3<br>Beta_F4–P4<br>Beta_F4–T4<br>Beta_P4–O2<br>Beta_T4–O2<br><br>Gamma_T3–O1<br>Gamma_P3–O1<br>Gamma_F3–T3<br>Gamma_F3–P3<br>Gamma_F4–P4<br>Gamma_F4–T4 |

| | | |
|---|---|---|
| | | Gamma_P4–O2<br>Gamma_T4–O2 |
| Single-channel features that are the ratio of powers | 8 channels × 3 ratios = 24 | ADR_T3–O1<br>ADR_P3–O1<br>ADR_F3–T3<br>ADR_F3–P3<br>ADR_F4–P4<br>ADR_F4–T4<br>ADR_P4–O2<br>ADR_T4–O2<br><br>BDR_T3–O1<br>BDR_P3–O1<br>BDR_F3–T3<br>BDR_F3–P3<br>BDR_F4–P4<br>BDR_F4–T4<br>BDR_P4–O2<br>BDR_T4–O2<br><br>ABDTR_T3–O1<br>ABDTR_P3–O1<br>ABDTR_F3–T3<br>ABDTR_F3–P3<br>ABDTR_F4–P4<br>ABDTR_F4–T4<br>ABDTR_P4–O2<br>ABDTR_T4–O2 |
| Single-channel features that do not use power | 8 channels × 2 = 16 | SEF90_T3–O1<br>SEF90_P3–O1<br>SEF90_F3–T3<br>SEF90_F3–P3<br>SEF90_F4–P4<br>SEF90_F4–T4<br>SEF90_P4–O2<br>SEF90_T4–O2<br><br>aEEG_T3–O1<br>aEEG_P3–O1<br>aEEG_F3–T3<br>aEEG_F3–P3<br>aEEG_F4–P4<br>aEEG_F4–T4<br>aEEG_P4–O2<br>aEEG_T4–O2 |
| Features that average over left-sided channels | 10 | Delta_1<br>Theta_1<br>Alpha_1<br>Beta_1<br>Gamma_1<br>ADR_1<br>BDR_1<br>ABDTR_1<br>SEF90_1<br>aEEG_1 |

| | | |
|---|---|---|
| Features that average over right-sided channels | 10 | Delta_2<br>Theta_2<br>Alpha_2<br>Beta_2<br>Gamma_2<br>ADR_2<br>BDR_2<br>ABDTR_2<br>SEF90_2<br>aEEG_2 |
| Features that average over all channels | 10 | Delta_0<br>Theta_0<br>Alpha_0<br>Beta_0<br>Gamma_0<br>ADR_0<br>BDR_0<br>ABDTR_0<br>SEF90_0<br>aEEG_0 |
| Asymmetry | 1 | Asymmetry |

**Table S2.** List of 65 qEEG features identified through feature selection. Note that at least one qEEG feature from each feature type is represented in the final selection.

| Feature type | Number of features | Description of features |
|---|---|---|
| Single-channel features that use power | 26 | Delta_T3–O1<br>Delta_P3–O1<br>Delta_F3–T3<br>Delta_F4–T4<br>Delta_P4–O2<br><br>Theta_T3–O1<br>Theta_P3–O1<br>Theta_F4–P4<br>Theta_F4–T4<br>Theta_P4–O2<br><br>Alpha_P3–O1<br>Alpha_F3–T3<br>Alpha_F4–T4<br>Alpha_P4–O2<br>Alpha_T4–O2<br><br>Beta_P3–O1<br>Beta_F3–T3<br>Beta_F3–P3<br>Beta_F4–T4<br>Beta_P4–O2<br>Beta_T4–O2<br><br>Gamma_P3–O1<br>Gamma_F3–T3<br>Gamma_F3–P3 |

| | | |
|---|---|---|
| | | Gamma_F4–P4<br>Gamma_P4–O2 |
| Single-channel features that are the ratio of powers | 9 | ADR_P3–O1<br>ADR_F4–T4<br>ADR_P4–O2<br>ADR_T4–O2<br><br>BDR_F4–P4<br>BDR_F4–T4<br><br>ABDTR_F3–T3<br>ABDTR_F3–P3<br>ABDTR_F4–P4 |
| Single-channel features that do not use power | 12 | SEF90_T3–O1<br>SEF90_P3–O1<br>SEF90_F3–P3<br>SEF90_F4–P4<br>SEF90_F4–T4<br>SEF90_T4–O2<br><br>aEEG_T3–O1<br>aEEG_P3–O1<br>aEEG_F3–T3<br>aEEG_F4–T4<br>aEEG_P4–O2<br>aEEG_T4–O2 |
| Features that average over left-sided channels | 8 | Delta_1<br>Beta_1<br>Gamma_1<br>ADR_1<br>BDR_1<br>ABDTR_1<br>SEF90_1<br>aEEG_1 |
| Features that average over right-sided channels | 7 | Delta_2<br>Alpha_2<br>Beta_2<br>Gamma_2<br>BDR_2<br>SEF90_2<br>aEEG_2 |
| Features that average over all channels | 2 | Alpha_0<br>Gamma_0 |
| Asymmetry | 1 | Asymmetry |

**Table S3**. Model performance using only qEEG features, as identified by AutoGluon for its top-performing model. Values in parentheses denote 95% confidence intervals.

| **Models** | **AUROC** | **AUPRC** | **Sens.** | **Spec.** | **Prec./PPV** | **NPV** |
|---|---|---|---|---|---|---|
| Ensemble-L3 | **0.96 (0.95–0.97)** | **0.44 (0.40–0.47)** | **0.84 (0.81–0.88)** | 0.96 (0.94–0.98) | 0.29 (0.23–0.33) | **0.998 (0.998–0.999)** |
